# Determinants of Antibody Levels and Protection against Omicron BQ.1/XBB Breakthrough Infection

**DOI:** 10.1101/2024.10.11.24315296

**Authors:** Carla Martín Pérez, Anna Ramírez-Morros, Alfons Jimenez, Marta Vidal, Edwards Pradenas, Diana Barrios, Mar Canyelles, Rocío Rubio, Inocencia Cuamba, Luis Izquierdo, Pere Santamaria, Benjamin Trinité, Josep Vidal-Alaball, Luis M. Molinos-Albert, Julià Blanco, Ruth Aguilar, Anna Ruiz-Comellas, Gemma Moncunill, Carlota Dobaño

## Abstract

The ongoing evolution of SARS-CoV-2, particularly through the emergence of new variants, continues to challenge our understanding of immune protection. While antibody levels correlate with protection against earlier variants like Alpha and Delta, their relationship with Omicron sub-variants remains unclear. To investigate the role of antibody levels and neutralizing activity in preventing breakthrough infections, we analyzed longitudinal SARS-CoV-2 humoral responses and neutralizing activity against the ancestral virus and major emerging variants in a well-characterized cohort of healthcare workers in Spain (N = 405). We found that antibody levels and neutralization titers are key indicators of protection against SARS-CoV-2, including the BQ.1 and XBB Omicron variants. Higher IgG and IgA levels were associated with protection over three 6-month follow-up periods sequentially dominated by BA.1, BA.2, BA.5, BQ.1, and XBB Omicron sub-variants, although the strength of the association between antibody levels and protection declined over time. Our findings demonstrate that binding antibody levels and neutralizing responses are a valid correlate of protection against more evasive BQ.1 and XBB Omicron variants, although the strength of this association declined over time. Additionally, our results underscore the importance of continuous monitoring and updating vaccination strategies to maintain effective protection against emerging SARS-CoV-2 variants.

## Introduction

The COVID-19 pandemic has led to unprecedented global health challenges since its emergence in late 2019. Vaccination has been the cornerstone of the public health response, significantly reducing severe disease, hospitalization, and mortality rates(1, 2). However, the continuous emergence of SARS-CoV-2 variants, particularly those classified as variants of concern (VOCs), has posed ongoing challenges to vaccine efficacy and public health strategies(3–6). Among these, the Omicron variant and its sub-lineages, including BA.1, BA.2, BA.5, BQ.1, XBB, BA.2.86, JN.1 and KP.3, have exhibited enhanced transmissibility and immune evasion capabilities(7–9), complicating efforts to control the spread of the virus and protect vulnerable populations.

Antibody-mediated immunity, particularly neutralizing antibodies, has been broadly demonstrated to play a crucial role in protecting against SARS-CoV-2 infection. Numerous studies have established that neutralizing capacity and antibody levels serve as correlates of protection against earlier variants such as Alpha and Delta(10–13). However, the relationship between antibody levels and protection against Omicron sub-variants remains less clear. While some studies have demonstrated a correlation between neutralizing capacity and antibody levels with protection against BA.1, BA.2(14–17), and BA.4/5(18) variants, others have found inconsistent results(19–21), especially for the subsequent sub-variants BQ.1 and XBB(22–25). These discrepancies highlight the need for a deeper understanding of the immune correlates of protection in the context of evolving SARS-CoV-2 variants. A strong association between binding antibody levels and neutralizing capacity against the ancestral strain in individuals with prior COVID-19 infection or vaccination has been reported by numerous studies(26–28). However, for neutralizing antibodies against Omicron subvariants, this relationship varies depending on previous exposure. While a weak correlation between antibody levels and neutralization has been observed in vaccinated non-infected participants(29, 30) and in those with two or fewer vaccine doses(31–33), a strong correlation has been noted in participants with three vaccine doses, particularly among those with breakthrough infections(33–35). Therefore, it is crucial to keep investigating this relationship in the context of emerging Omicron variants.

Hybrid immunity, resulting from a combination of vaccination and natural infection, has been shown to induce higher and more durable antibody responses compared to vaccination or infection alone(36–40). Nevertheless, the extent to which hybrid immunity confers protection against newer Omicron sub-variants is not fully understood(11, 18, 22, 38, 41). Additionally, the impact of prior infections and the timing of these infections on the protection against these sub-variants warrants further investigation.

This study aimed to address these knowledge gaps by investigating longer-term anti-SARS-CoV-2 antibody dynamics up to three years after the start of the pandemic, spanning three 6-month follow-up periods in a well-characterized prospective cohort. We evaluated antibody levels and neutralizing activity as correlates of protection during periods dominated by the BA.1, BA.2, BA.5, BQ.1, and XBB Omicron sub-variants. To detect potential asymptomatic infections alongside reported symptomatic cases, we also leveraged serological data obtained over 11 longitudinal visits. Additionally, we examined the impact of prior SARS-CoV-2 infection, hybrid immunity, booster immunization (third and fourth dose) with COVID-19 mRNA vaccines, as well as clinical and demographic factors, on antibody levels. Specifically, we assessed how these factors influence protection against breakthrough infections, considering both symptomatic and asymptomatic cases.

## Results

### Cohort and study design

The CovidCatCentral (CCC) cohort consists of two groups of primary healthcare workers (HCWs) from three primary care counties in Barcelona, Spain(42) (**Fig. 1**). The first group includes 247 individuals enrolled from September 2020 to January 2021 who had experienced symptomatic SARS-CoV-2 infections prior to recruitment. The second group comprises 200 naïve HCWs recruited after receiving full primary vaccination (March – April 2021). The first group was visited at 11 timepoints (T) while the second was visited at 7 timepoints, between September 2020 and June 2023 (n at T9: 377, n at T10: 392, n at T11: 346) (**Table. 1**). Most of the study participants were female (83.8% at T9, 85.2% at T10, and 86.4% at T11). The mean age of the participants was 48.6 years (standard deviation [SD] 10.7) at T9, 50.0 years (SD 10.7) at T10, and 50.0 years (SD 10.4) at T11. Additionally, 66.8% of participants at T9, 66.3% at T10, and 70.2% at T11, had underlying comorbidities.

**Figure 1.**
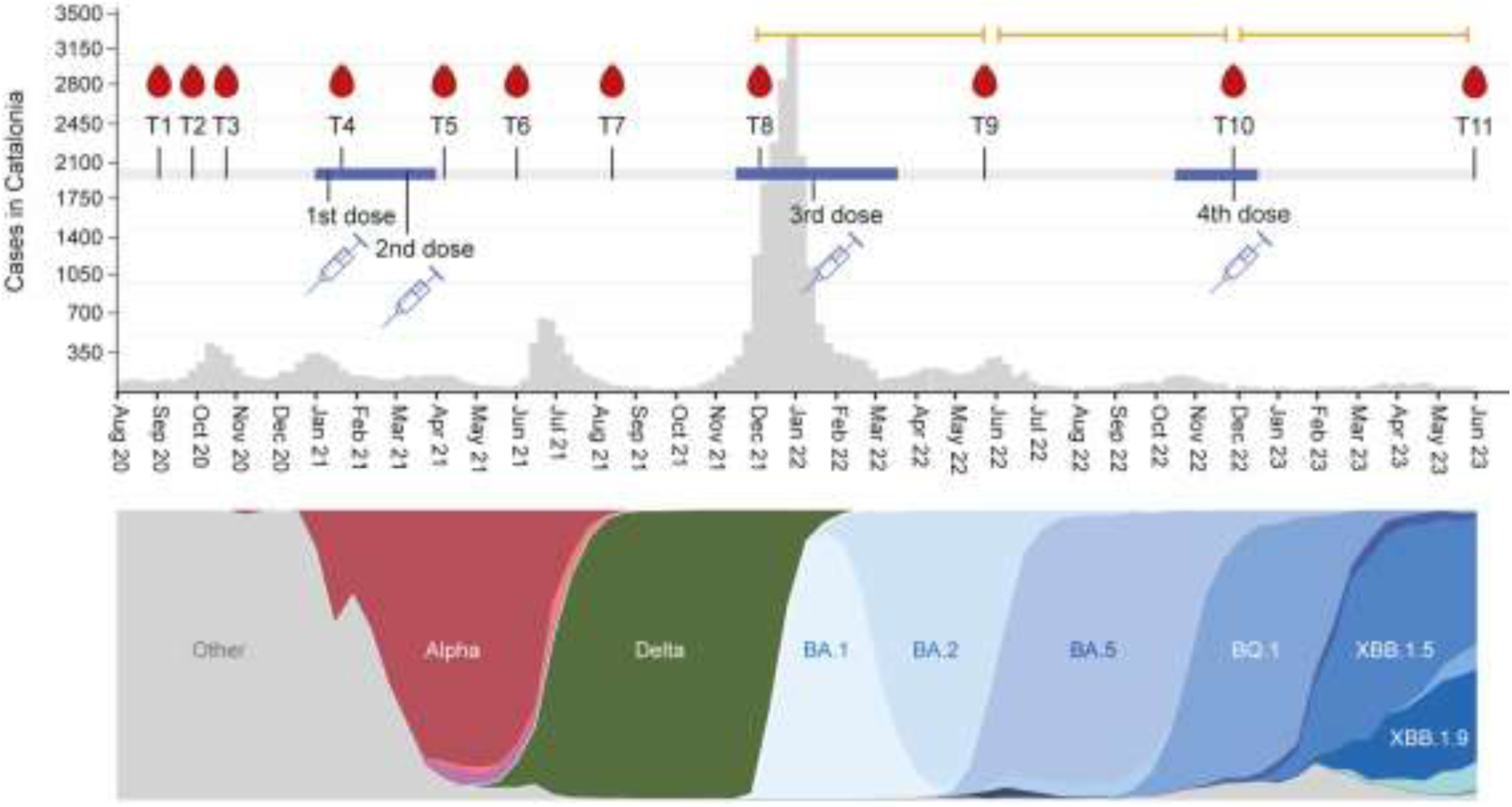
CCC study sample collection and vaccination timepoints, SARS-CoV-2 cases in Catalonia, and main SARS-CoV-2 lineages circulating in Spain over the study period. The grey plot represents the number of SARS-CoV-2 cases at a given time in Catalonia according to the official data available from IDESCAT (Statistical Institute of Catalonia, https://www.idescat.cat/dades/covid19/?lang=es (accessed on 03 July 2024)). The bottom timeline depicts the main SARS-CoV-2 lineages circulating in Spain at those time intervals according to GISAID. The orange lines represent the three follow-up periods (6 months each) for breakthrough infections in this study. T: Timepoint.

### SARS-CoV-2 infections and COVID-19 vaccination

Symptomatic SARS-CoV-2 infections were detected by real-time reverse transcription polymerase chain reaction (rRT-PCR) and/or rapid diagnostic test (RDT). To identify potential undiagnosed COVID-19 infections (asymptomatic infections), we used already available serology data and employed a fold-change analysis of antibody levels to spike (S) and nucleocapsid (N) antigens across consecutive study timepoints. We identified 2, 2, 14, 8, 10, 30, 39, and 55 asymptomatic infections within the intervals: T3-T4, T4-T5, T5-T6, T6-T7, T7-T8, T8-T9, T9-T10, and T10-T11, respectively. Using this method, we were able to detect at least 75% of the diagnosed infections in each interval, which indicates we had a minimum of 75% sensitivity to detect the undiagnosed infections (**Suppl Table 1**).

At T9 (May 2022, n = 377) (**Suppl Table 2**), 333 participants had received the primary series of vaccination, and 245 had received the 1^st^ booster dose. The median time since last vaccination was 157 days (27 interquartile range [IQR]). Up to T9, 79.6% of the participants (n = 300) had been previously infected by SARS-CoV-2 according to rRT-PCR, RDT, or serology data, and 20.4% (n = 77) had no evidence of infection. The median time since last infection was 204 days (666.5 IQR). At T10 (November - December 2022, n = 392) (**Suppl Table 3**), 89% of participants (n = 350) had received the primary series of vaccination, 67% (n = 264) had received a 3^rd^ dose (1^st^ booster), and 10% (n = 41) had received the 2^nd^ booster dose. The median time since last vaccination was 342.5 days (37 IQR). Up to T10, 89% of the participants (n= 349) had been previously infected by SARS-CoV-2, and 10.9% (n = 43) had no evidence of infection. The median time since last infection was 279 days (251 IQR). At T11 (May - June 2023, n = 346) (**Table 1**), 14 participants had not received any vaccine doses (4%), 308 participants had received the primary series of vaccination (89%), 241 had received a 3^rd^ dose (70%), and 72 had received the 2^nd^ booster dose (21%). The median time since last vaccination was 514 days (234.5 IQR). Up to T11, 94.2% of the participants (n= 326) had been previously infected by SARS-CoV-2, and 5.8% (n = 20) had no evidence of infection but were vaccinated with 3 or 4 doses. The median time since last infection was 360.5 days (219.25 IQR).

**Table 1.** Characteristics of study participants at T11.

| | | N or mean ± SD |
| --- | --- | --- |
| Age | | 50 ± 10.4 |
| Sex | Male | 47 |
|  | Female | 299 |
| Comorbidities | Yes | 243 |
|  | No | 103 |
| Smoking | Yes | 144 |
|  | No | 202 |
| Vaccine doses | 0 | 14 |
|  | 1 | 24 |
|  | 2 | 67 |
|  | 3 | 169 |
|  | 4 | 72 |
| Number of infections* | 0 | 20 |
|  | 1 | 126 |
|  | 2 | 147 |
|  | 3 | 49 |
|  | 4 | 4 |
| First dose type | Astrazeneca | 5 |
|  | Janssen | 1 |
|  | Moderna | 4 |
|  | Pfizer | 300 |
|  | NA | 9 |
| Second dose type | Astrazeneca | 3 |
|  | Moderna | 12 |
|  | Pfizer | 271 |
|  | NA | 11 |
| Third dose type | Moderna | 105 |
|  | Pfizer | 95 |
|  | NA | 27 |
| Fourth dose type | Pfizer | 66 |
|  | NA | 6 |
| First exposure type | Infection | 185 |
|  | Vaccination | 161 |

\* Includes symptomatic and asymptomatic infections

Among the vaccinated participants, 141 (36.9%) had vaccine breakthroughs between T8 – T9 (113 diagnosed and 28 detected by serology), 128 (35.3%) had vaccine breakthroughs between T9–T10 (92 diagnosed and 36 detected by serology), and 89 (23.7%) had vaccine breakthroughs between T10 – T11 (54 diagnosed and 35 detected by serology).

### Recent infection, hybrid immunity, and booster doses are associated with higher IgG antibody levels

At T9, those with hybrid immunity exhibited higher IgG levels against all measured antigens, corresponding to the Wuhan strain (Beta coefficient [β] anti-S IgG 21.9%, 95% confidence interval [CI]: 52.0-89.6) (**Fig. 2, Suppl Fig. 1**). Similarly, the number of prior infections correlated with increased IgG levels against all Wuhan antigens. The number of vaccine doses received was associated with elevated IgG levels against S, S1, and receptor binding domain (RBD) (β anti-S IgG 67.5%, 95% CI: 51.5-85.2). Receiving three doses, as opposed to two, was linked to higher IgG levels against spike antigens (β anti-S IgG 52.5%, 95% CI: 22.1-90.3). In addition, age was positively associated with higher levels of IgG to S2, and individuals with comorbidities had higher levels of IgG against S antigens.

**Figure 2.**
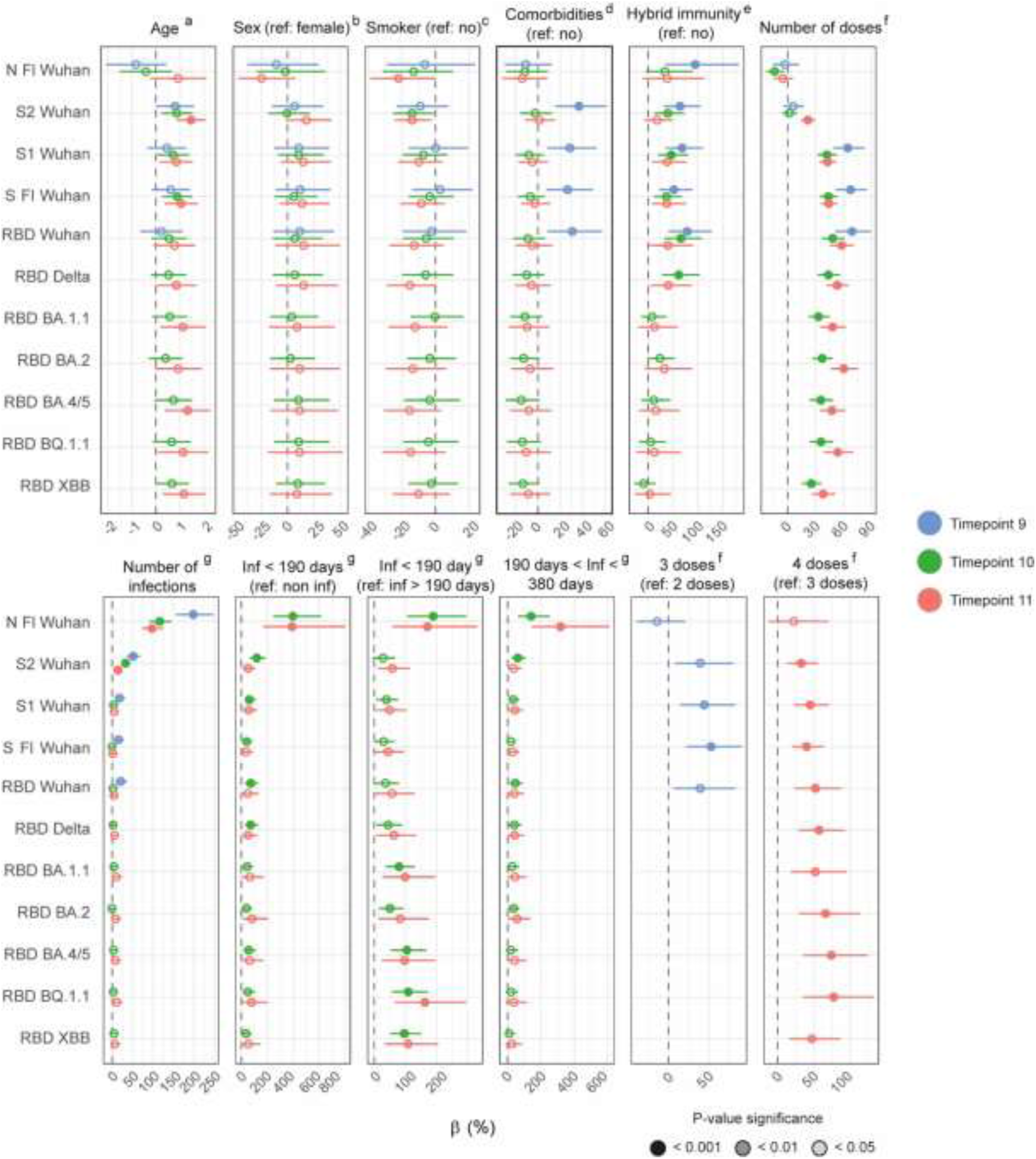
Association of clinic-demographic factors with IgG antibody levels at T9 (red), T10 (green) and T11 (blue) using multivariable linear regression models in vaccinated individuals. Beta (β) and CI values have been transformed to a percentage for an easier interpretation. The color inside of the dots represents the P value after adjustment for multiple testing by Benjamini-Hochberg, where dark color represents < 0.001, intermediate color < 0.01, light color < 0.05, and white non-significant. ^a^ Adjusted by sex. ^b^ Adjusted by age. ^c^ Adjusted by age and sex. ^d^ Adjusted by age, sex, smoking, number of doses and number of infections. ^e^ Adjusted by age, sex, comorbidities, first exposure type, time since last exposure, number of doses and number of infections. ^f^ Adjusted by age, sex, comorbidities, smoking, and number of infections. ^g^ Adjusted by age, sex, comorbidities, smoking, and number of doses. Full-length (Fl), Nucleocapsid (N), Receptor-binding domain (RBD), Spike (S).

Having hybrid immunity was associated with higher IgG levels to S and S1 from Wuhan and RBD from Wuhan and Delta variants at T10 (β anti-S Fl IgG 36.9%, 95% CI: 11.7-67.9) and T11 (β anti-S Fl IgG 37.9%, 95% CI: 8.03-76.2), as well as Wuhan S2 at T10. However, at both timepoints, hybrid immunity did not correlate with IgG levels to Omicron antigens (**Fig. 2, Suppl Fig. 1**).

The number of vaccine doses received were positively associated with IgG levels against all S antigens at both T10 (β anti-S Fl IgG 43.7%, 95% CI: 34.1-53.9) and T11 (β anti-S Fl IgG 44.2%, 95% CI: 35.4-53.6), with the exception of S2 from Wuhan at T10. (**Fig. 2, Suppl Fig. 1**). Notably, at T10, the number of doses was negatively associated with IgG levels to N. By T11, receiving four doses compared to three was linked to higher IgG levels against all antigens except N (β anti-S Fl IgG 45.5%, 95% CI: 20.9-75.0).

The number of prior infections consistently showed a positive association with IgG levels to S2 and N proteins at all three timepoints (**Fig. 2, Suppl Fig. 1**). Additionally, at T10, a recent infection (within 190 days) was linked to higher IgG levels against all antigens compared to no infection (β anti-S1 IgG 79.1%, 95% CI: 46.3-116.9), and higher IgG levels to RBD from Omicron and Delta variants, as well as against Wuhan S1 and N, compared to having an infection more than 190 days prior (β anti-S1 IgG 37.6%, 95% CI: 13.8-66.4). At T11, a recent infection similarly correlated with higher IgG levels against Delta RBD, BA.2 RBD, BQ.1.1 RBD, and Wuhan S1, S2, and N, as compared to no infection (β anti-S1 IgG 59.95%, 95% CI: 23.1-107.8), whereas infections occurring within 190 days associated with higher IgG levels to all antigens except Wuhan RBD compared to infections more than 190 days prior (β anti-S1 IgG 19.1%, 95% CI: 2.99-38.6). Infections between 190- and 380-days prior were associated with elevated IgG levels against Delta RBD, BA.2 RBD, and Wuhan S1 and N at both T10 (β anti-S1 IgG 91.7%, 95% CI: 8.80-237.9) and T11 (β anti-S1 IgG 32.6%, 95% CI: 5.15-67.2), as well as IgG to RBD and S2 at T10, compared to no infection (**Fig. 2, Suppl Fig. 1**).

Age was consistently positively associated with IgG levels to S, S1, and S2 antigens at both T10 and T11, with additional associations observed with all Omicron RBDs except BA.2 at T11 (**Fig. 2, Suppl Fig. 1**). Lastly, smoking was inversely associated with IgG levels to S2 at T10 and with S2 and N at T11.

### Having a recent infection is associated with lower risk of Omicron breakthrough infection

The number of previous infections was positively associated with protection against symptomatic infections during the T9–T10 and T10–T11 six-month periods in multivariable Cox models (T9–T10 hazard ratio [HR], 0.55; 95% CI, 0.40-0.76; T10–T11 HR, 0.56; 95% CI, 0.35-0.91) (**Fig. 3a, Suppl Fig. 2a**). Similarly, a higher number of previous infections was associated with protection against all infections (symptomatic plus asymptomatic) in both periods according to multivariable logistic regression models (T9–T10 odds ratio [OR], -54.5%; 95% CI, -31.4 to -70.4; T10–T11 OR, -56.7%; 95% CI, -34.7 to -71.9) (**Fig. 3b, Suppl Fig. 2b**). Furthermore, having a recent infection within the previous 190 days was associated with protection against symptomatic infections over both the T9–T10 and T10–T11 periods in multivariable Cox models (T9–T10 HR, 0.15; 95% CI, 0.07-0.31; T10–T11 HR, 0.007; 95% CI, 0.01-0.46) (**Fig. 3a, Suppl Fig. 2a**). This protective effect was also observed for all infections (T9–T10 OR, -79%; 95% CI, -53.9 to -90.9; T10–T11 OR, -83.8%; 95% CI, -54.5 to -94.6) (**Fig. 3b, Suppl Fig. 2b**). In unadjusted models, hybrid immunity was associated with protection in T9 –T10 (HR, 0.4; 95% CI, 0.61 to 0.26; OR, -54%; 95% CI, -21.9 to -73.9) and T10 – T11 (HR, 0.38; 95% CI, 0.92 to 0.15; OR, -67.3%; 95% CI, -29.4 to -85.0) periods, however, this association was lost after adjusting for the number of exposures and time since the last exposure (**Fig. 3a,b, Suppl Fig. 2a,b**).

**Figure 3.**
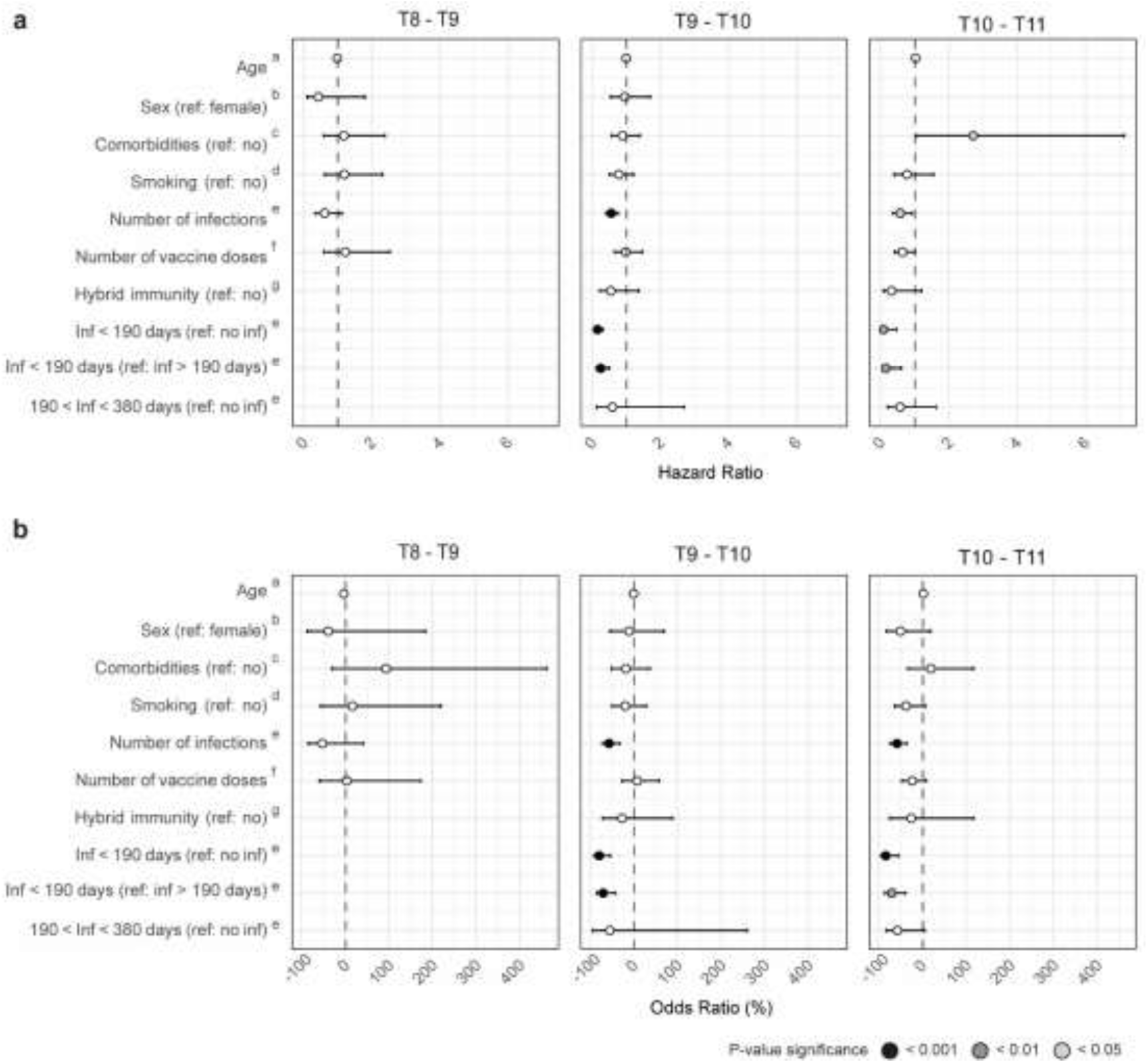
Association of clinic-demographic factors with protection against symptomatic (a), and symptomatic and asymptomatic infections (b) using multivariable Cox and Logistic regression models, respectively. In b, odds ratio and CI values have been transformed to a percentage for an easier interpretation. The color of the dots represents the P value, where black represents < 0.001, dark grey < 0.01, light grey < 0.05, and white non-significant. ^a^ Adjusted by sex. ^b^ Adjusted by age. ^c^ Adjusted by smoking, age and sex. ^d^ Adjusted by age and sex. ^e^ Adjusted by age, sex, comorbidities, smoking, and number of doses. ^f^ Adjusted by age, sex, comorbidities, smoking, and number of infections. ^g^ Adjusted by age, sex, comorbidities, first exposure type, time since last exposure, number of doses and number of infections.

On the other hand, comorbidities were associated with a higher risk of symptomatic breakthrough infections during the T10–T11 period (HR, 2.7; 95% CI, 1.02-7.13) (**Fig. 3a, Suppl Fig. 2a**). In contrast, age, sex, the number of vaccine doses, and having an infection between 190- and 380-days prior were not found to be associated with protection during any of the examined periods.

### Antibody levels to previous VoC correlate with protection against Omicron breakthrough infection

We evaluated the relationship between IgG and IgA levels and protective immunity across the three consecutive time periods (T8–T9, T9–T10, and T10–T11). We measured antibodies to the Wuhan strain: RBD, S Fl, S2 and N for all periods, and S1 for T9–T10 and T10–T11. Additionally, during the T8–T9 period, we measured antibodies targeting the RBD from Delta, Alpha, Beta, and Gamma variants, and during the T10-T11 period, antibodies to RBD from Omicron sub-variants (BA.1.1, BA.2, BA.4/5, BQ.1.1, and XBB). We found an association between antibody levels and protection against symptomatic infections for all tested antibodies except IgG to S2 Wuhan at T9-T10 and T10-T11, and IgA to S2 and S Wuhan at T9-T10 (**Fig. 4a**). Similarly, we found an association between antibody levels and protection against symptomatic plus asymptomatic infections for all antibodies except IgA to any antigen at T9-T10, and IgG to S2, RBD Wuhan and RBD Delta at T10-T11 (**Suppl Fig. 3**).

**Figure 4.**
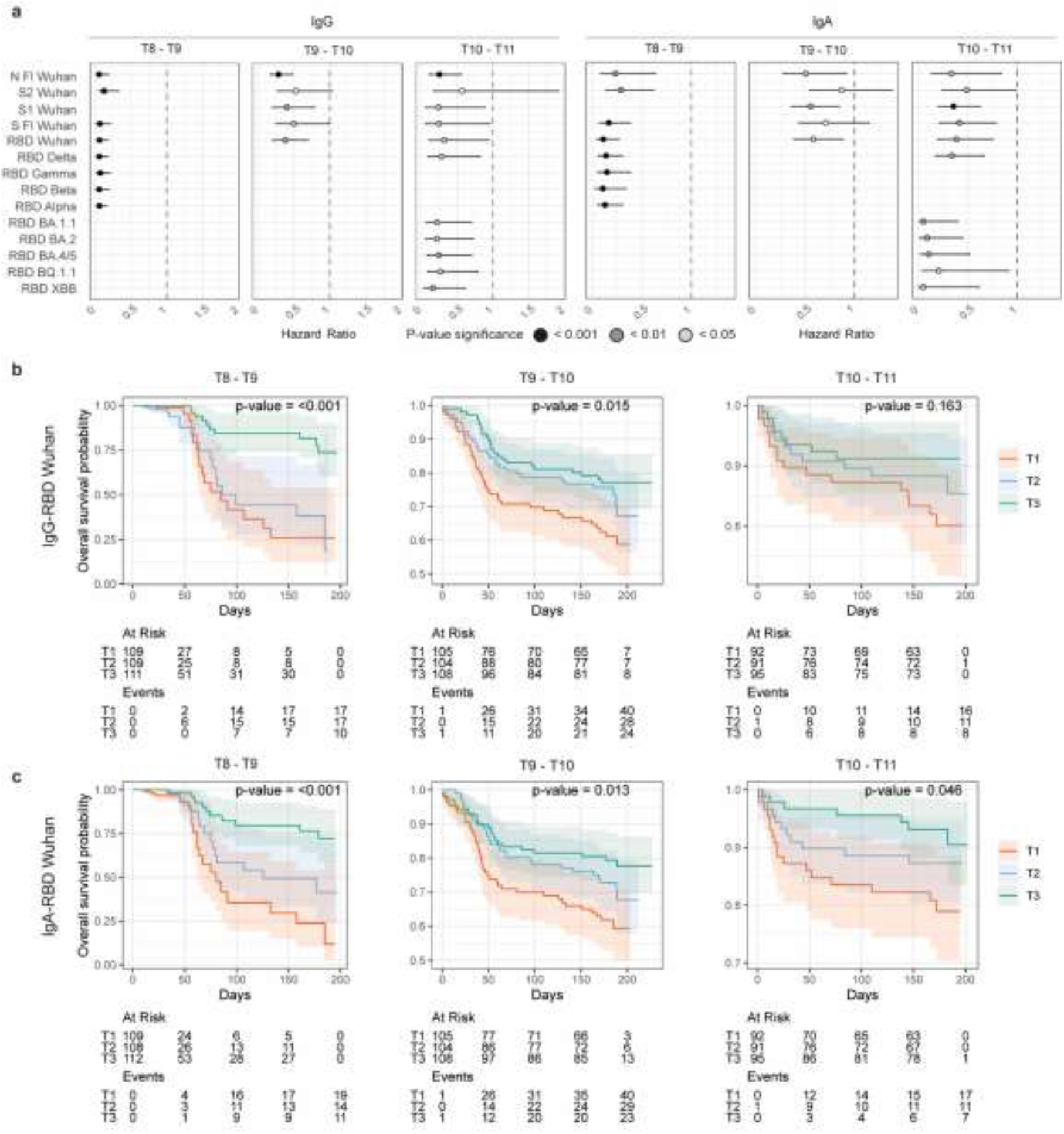
Association of antibody levels with protection against symptomatic breakthrough infections in vaccinated individuals at T8 – T9, T9 – T10 and T10 – T11 periods. **a**. Forest plot of multivariable Cox Regression models. Models were adjusted by age, comorbidities, hybrid immunity, number of infections, number of doses, sex, smoking, and time since last exposure. The color of the dots represents the P value, where black represents < 0.001, dark grey < 0.01, light grey < 0.05, and white non-significant. **b, c**. Kaplan–Meier survival curves of risk of breakthrough infection by tertiles of anti-RBD IgG (b) and IgA levels (c). Tertile T1 corresponds to the lowest antibody levels, whereas T3 denotes the highest. Shaded areas represent the 95% confidence intervals. Full-length (Fl), Nucleocapsid (N), Receptor-binding domain (RBD), Spike (S). Kaplan–Meier curves were compared using the log-rank test.

IgG and IgA antibody levels to RBD Wuhan were stratified into tertiles, and survival curves plotted for each level (**Fig. 4b**). Higher levels of IgG to RBD were associated with increased protection against symptomatic infection during the T8–T9 and T9–T10 periods, but not during the T10–T11 period. Similarly, higher levels of IgA-RBD were associated with protection across all three periods. Nevertheless, the differences between tertiles were most pronounced in the T8–T9 period compared to subsequent periods.

### Changes in the correlation of antibodies with protection over time

To investigate how antibody protective effect varied across the three time-periods, we assessed the interaction between the time-period and antibody levels using multivariable logistic regression and Cox models (**Suppl Table 4** and **5**, respectively). The association between antibody levels and protection against symptomatic and asymptomatic infections was stronger during the T8–T9 than the T9–T10 period for IgG and IgA to RBD and S (**Suppl Table 4**). Similarly, for IgG to RBD from the Delta variant, and RBD, S, S2, and N from Wuhan, the association was stronger during the T8–T9 than the T10–T11 period (**Suppl Table 4**). The association between antibody levels and protection against only symptomatic infections was also stronger during the T8–T9 compared to the T9–T10 period for IgG to RBD, S2, S and N, and for IgA to RBD, S, and S2 (**Suppl Table 5**). Similarly, for IgG to RBD from Delta variant, and RBD, S, and S2 from Wuhan, the association was stronger during the T8–T9 than the T10–T11 period (**Suppl Table 5**). No significant differences were found between the T9–T10 and T10–T11 periods.

Notably, when comparing the protective effect of antibody levels against Wuhan RBD during the T8–T9 period with the protective effect of antibody levels against Omicron RBDs during the T10– T11 period, we observed that the association with protection against symptomatic infections was generally stronger for IgG to Wuhan RBD at T8 than for IgG to Omicron RBD at T10 (**Suppl Table 6a**). Similarly, the association with protection against all infections was stronger for IgG against Wuhan RBD at T8 than for IgG against RBD from all tested Omicron sub-variants (BA.1.1, BA.2, BA.4/5, BQ.1.1, XBB) at T10 (**Suppl Table 6b, Fig. 4a, Suppl Fig. 3a**). Regarding protection against symptomatic infections, the association of IgG against Wuhan RBD was stronger than that of BQ.1.1 and BA.4/5 RBDs (**Suppl Table 6a, Fig. 4a, Suppl Fig. 3a**). This suggests that the decline in the protective effect of antibody levels over time is not due to lower antibody levels against the newer Omicron variants but due to a reduced antibody functionality.

### Neutralizing capacity correlates with protection against Omicron breakthrough infection

We found a positive correlation between antibody levels to S antigens and plasma neutralizing activity at T9 (**Fig. 5a**) and T11 (**Suppl Fig. 4**), particularly for the anti-Omicron antibodies at T11. When we analyzed the neutralizing activity against the Wuhan, BA.1, BA.4/5, and BQ.1.1 variants at T9 and its relation to protection against symptomatic or all infections during the T9–T10 period in a subset of participants (n = 113) (**Fig. 5, Suppl Table 7**), we found a positive association between neutralizing activity to Wuhan (D614G) and Omicron BA.4/5 variants and protection against symptomatic infections in a multivariable Cox regression model (**Fig. 5b, Suppl Table 7a**). Similarly, higher neutralizing activity against Wuhan and Omicron BA.1 and BA.4/5, were associated with protection against symptomatic and asymptomatic infections in a multivariable Logistic regression model (**Fig. 5c, Suppl Table 7b**).

**Figure 5.**
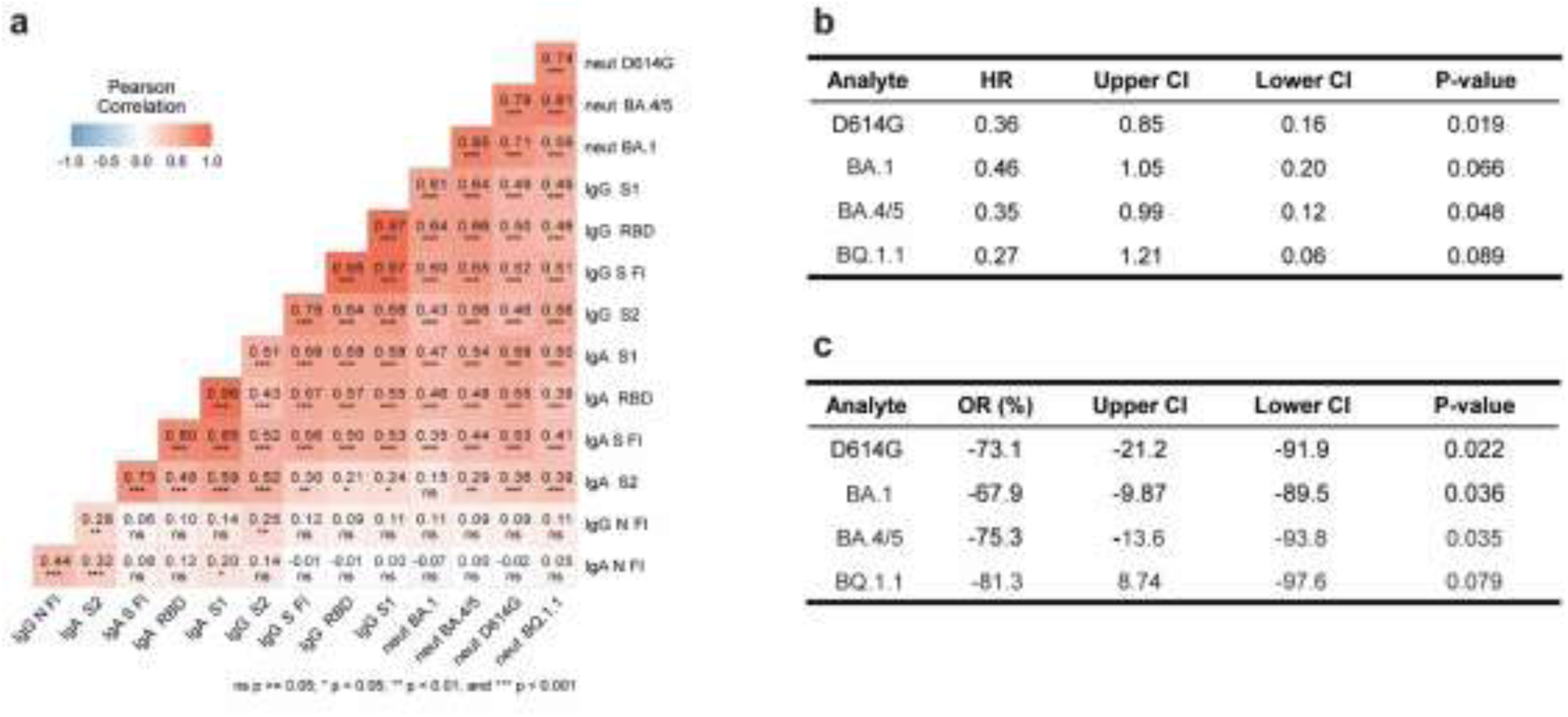
Association of antibody neutralizing capacity with protection against symptomatic and asymptomatic infections between T9 and T10 (Jun 2022 – Dec 2022) in vaccinated individuals. **a**. Correlation between antibody levels and plasma neutralization capacity at T9. Pearson correlation R values and correlation p-values (* p < 0.05, ** p < 0.01, *** p < 0.001, ns) are shown in the heatmap. **b**. Summary of multivariable Cox regression models assessing the association between plasma neutralizing capacity at T9 and protection against symptomatic infections over the T9 – T10 period. **c**. Summary of multivariable Logistic regression models assessing the association between plasma neutralizing capacity at T9 and protection against symptomatic and asymptomatic infections over the T9 – T10 period. Odds ratio and CI values have been transformed to a percentage for an easier interpretation. The D614G variant corresponds to the original Wuhan strain of the virus. Cox and Logistic regression models were adjusted by age, comorbidities, hybrid immunity, number of infections, number of doses, sex, smoking, and time since last exposure.

## Discussion

In this longitudinal study, we evaluated SARS-CoV-2 antibody levels and neutralizing activity as correlates of protection against both symptomatic and asymptomatic infections during COVID-19 pandemic periods dominated by the BA.1, BA.2, BA.5, BQ.1, and XBB Omicron sub-variants. Our findings indicate that antibody levels correlate with plasma neutralizing activity and both associate with protection against later as well as earlier Omicron variants, with the protective effect being more prominent initially and potentially decreasing over time.

Specifically, IgG and IgA levels to Wuhan, Delta, and Omicron variants were associated with protection against SARS-CoV-2 across all three follow-up periods. However, the strength of the association between higher antibody levels and lower COVID-19 risk decreased in the T9-T10 (when BA.5 variant prevailed) and T10-T11 (when BQ.1 and XBB variants were prevalent) periods, compared to the T8-T9 period (when BA.1 and BA.2 variants prevailed). Furthermore, the association between anti-Wuhan IgG levels with protection during the T8-T9 period was stronger than that of anti-Omicron IgG levels during the T10-T11 period. Our findings suggest that more effective antibodies may be required to protect against infections with BA.5, BQ.1, and XBB Omicron variants compared to BA.1/BA.2 infections. The decline in the protective effect of antibody levels over time may be linked to a shift in the proportion of IgG subclasses with varying effector functions, specifically a shift towards IgG4, which has been previously associated with mRNA vaccination(43, 44). Furthermore, this decline may also result from the enhanced immune escape of newer Omicron variants, leading to reduced antibody functionality. This is consistent with previous studies reporting enhanced immune evasion of BQ.1.1 and XBB as compared to earlier Omicron variants, including BA.5 and BA.2(7–9, 45, 46). Moreover, we found that at T9 and T11, anti-Wuhan and anti-Omicron antibody levels had a strong correlation with neutralizing activity. This indicates that anti-Omicron antibody levels can still serve as a surrogate for neutralizing antibody efficacy, highlighting the utility of monitoring anti-S IgG binding antibodies to infer neutralizing capacity. Nevertheless, this relationship should be closely observed as new variants emerge.

Multiple studies have previously established that antibody levels are a correlate of protection against infections involving Alpha and Delta variants(11–13). However, this correlation has been less clear for Omicron variants BA.1, BA.2, and BA.5, and particularly for BQ.1 and XBB. Several studies found that higher IgG to S and neutralizing activity were linked to protection against BA.1 and BA.2 infection(15–17). However, other studies found no consistent association, with some reporting a link between antibody levels and protection against BA.1 but not BA.2(19–21). Regarding BA.5, BQ.1, and XBB variants, some research suggests higher IgG levels increase protection against BA.4/5 infection(18, 22, 47) and XBB infection(24, 25), but other studies found no correlation during BA.5 and XBB waves(22, 23). These conflicting results may be due to variations in study design, sample size, duration of follow-up, previous infection status of the participants and differences in neutralization and antibody level measurement assays. Here, identifying asymptomatic infections among undiagnosed participants by tracking changes in antibody levels may have enhanced the sensitivity of our correlates of protection analysis.

Our findings also indicate that hybrid immunity was consistently associated with higher anti-Wuhan and anti-Delta IgG levels at all timepoints studied. However, this association did not extend to IgG levels against Omicron variants. Contrary to previous studies, which reported no sustained association between hybrid immunity and elevated antibody levels following a booster dose or a third exposure event(36, 38), our analysis still detected this association even when restricted to participants who had received three vaccine doses. Despite the observed higher IgG levels to the original Wuhan strain and Delta variant, hybrid immunity did not confer enhanced protection against Omicron breakthrough infections in models that included number of exposures and time since last exposure as covariates. This could be attributed to the lack of higher anti-Omicron IgG levels in individuals with hybrid immunity. Data suggest that while hybrid immunity boosts antibody levels against earlier strains, it may not significantly enhance immunity against newer Omicron variants independent of the number of exposures and time since last exposure. In previous studies, hybrid immunity has been shown to have a protective effect against Omicron infection(18, 22, 41, 48). However, these studies did not adjust for the number of exposures or the time since the last exposure.

The number of vaccine doses was consistently associated with higher IgG levels against all S antigens and variants at the three timepoints studied. In contrast, the number of infections was associated with elevated IgG levels to S Fl, RBD, and S1 only at T9, but not at T10 or T11, although the number of infections was consistently associated with anti-S2 and anti-N IgG levels across all timepoints. Having a recent infection (within the past year or the past six months) was linked to higher IgG levels at T10 and T11. Importantly, having recent infection within the past six months was associated with increased protection on T9-T10 and T10-T11 periods. However, despite the association between previous infection within the past year and higher IgG levels, this did not translate into increased protection during the T9-T10 or T10-T11 periods. The strong correlation between the time since the last infection and the variant involved makes it challenging to desintangle the individual impact of each factor. However, a previous study suggests that protection is primarily influenced by the variant of the prior infection, rather than the time elapsed since the last infection(18). In our study, infections occurring 6 to 12 months prior to T9 were most likely caused by the Delta variant, while those within the past 6 months were predominantly BA.1 and BA.2. By T10, infections 6 to 12 months prior were most likely caused by Omicron BA.1 and BA.2, whereas those within the past 6 months were predominantly BA.5. This indicates that although both groups of infections during the T10 period were Omicron, only the more recent infections (within 6 months) were associated with increased protection. This suggests that the closeness to the infection, possibly due to higher levels of specific antibodies that decay over time, plays an important role in conferring protection. The number of infections also correlated with protection during the T9-T10 period, aligning with the observed higher IgG levels at T9. Moreover, although the number of infections was not associated with higher IgG levels at T10, it was still associated to protection during the T10-T11 period, suggesting that other components of the immune response, including cellular immunity, could be involved.

Our study has limitations. First, our cohort primarily consisted of young adult women from the HCW population, which may not fully represent the general population. Second, we did not have specific data on the viral strains responsible for breakthrough infections. However, our analysis was conducted during periods when sequencing data from Spain indicated a dominance of BA.1 and BA.2 (T8-T9), BA.5 (T9-T10), or BQ.1 and XBB (T10-T11)(49). Third, we did not assess neutralizing responses for all participants, although the correlation between antibody levels and neutralizing activity was very high, especially for antibodies to Omicron. Additionally, we measured plasma rather than mucosal IgA, which might be more relevant for immunity against infection.

In summary, our study reveals that antibody levels and neutralizing activity remain important correlates of protection to SARS-CoV-2, even against Omicron variants like BQ.1 and XBB. Our findings indicate that while higher anti-S IgG levels are associated with increased protection during periods dominated by BA.1, BA.2, BA.5, BQ.1 and XBB variants, the efficacy of these antibodies declines over time. Importantly, recent infections provide significant protection, emphasizing the role of both the timing and the variant of prior infections in conferring protection. These results extend previous findings by showing that binding antibody levels are a valid correlate of protection against BQ.1 and XBB Omicron variants and highlight the importance of serological markers in assessing infection risk and informing booster vaccination policies. Continuous monitoring of antibody dynamics against new JN.1 and KP.2 variants and timely updates to vaccination strategies are crucial to maintain robust protection against evolving SARS-CoV-2 variants.

## Materials and Methods

### Sex as biological variable

Our study included both males and females, and no sex-based differences were observed in the findings.

### Study design and setting

The CovidCatCentral cohort comprises two groups of primary HCW recruited from three primary care counties in Barcelona, Spain. The first group consists of individuals recruited during the first wave of the COVID-19 pandemic (March–April 2020, n = 247) who had a symptomatic SARS-CoV-2 infection confirmed by rRT-PCR and/or RDT. All HCWs with COVID-19 were invited to participate. Questionnaires and venous blood samples were collected at eleven cross-sectional surveys up to the end of June 2023.

The second group includes naïve HCWs recruited from March–April 2021 after completing full primary vaccination (n = 200). This group was selected to have similar characteristics (age, sex, professional category, smoking habits) to the pre-exposed group. These HCWs were visited at seven cross-sectional surveys, with venous blood samples collected up to the end of June 2023. Demographic and clinical data were collected at baseline and during follow-up visits through telephone interviews and electronic questionnaires conducted by study physicians and nurses. The recorded information included smoking status, comorbidities, confirmed SARS-CoV-2 infections and COVID-19 symptoms, vaccination type and dates, and adverse effects. Diagnosed infections (symptomatic) were identified by passive case detection, while undiagnosed infections (asymptomatic) were identified by serology using the method described below.

### Quantification of antibodies to SARS-CoV-2

We measured IgA and IgG level (median fluorescence intensity, MFI) to the full-length (Fl) SARS-CoV-2 N and S antigens, the S subregions S1 and S2, and the RBD from different variants (Wuhan, Alpha, Beta, Delta, Gamma, BA.1, BA.2, BA.4/5, BQ.1.1, and XBB), by quantitative suspension array technology assays (xMAP, Luminex), following a previously described protocol(50). A nucleotide fragment encoding the ancestral N FL, followed by a 6xHis-tag, was cloned into pET22b expression vector, transformed in *E. coli* BL21 DE3, induced with IPTG, and purified by affinity chromatography using HisTrap columns, and controlled for purity by SDS-PAGE and Coomassie staining(51). The ancestral S and the RBD proteins were fused with C-terminal 6xHis and StrepTag sequences and purified from the supernatant of lentiviral-transduced CHO-S cells cultured under a fed-batch system(52). The generation of RBD proteins from the Alpha, Beta, and Gamma variants has been detailed in a previous study(42). Codon-optimized nucleotide fragments encoding the RBD variants (Delta, BA.1, BA.2, BA.4/5, BQ.1.1, and XBB) were synthesized and cloned into pcDNA3.1/Zeo (+) expression vector (Thermo Fisher Scientific)(53). Recombinant proteins were produced by transient transfection of exponentially growing Freestyle TM 293-F suspension cells (Thermo Fisher Scientific) using the polyethylenimine (PEI)- precipitation method. Proteins were purified from culture supernatants by high-performance chromatography using the Ni Sepharose® Excel Resin (GE Healthcare), according to manufacturer’s instructions, dialyzed against PBS using Slide-A-Lyzer® dialysis cassettes (Thermo Fisher Scientific), quantified using NanoDrop TM One instrument (Thermo Fisher Scientific), and controlled for purity by SDS-PAGE using NuPAGE 3-12% Bis-tris gels (Life Technologies). Plasma samples were tested at a 1:500 dilution for the two isotypes and additionally at a 1:5000 dilution for IgG to prevent saturation of anti-S levels in vaccinated participants. To quantify IgA, samples and controls were pretreated with anti-human IgG (Gullsorb) at 1:10 dilution, to avoid IgG interferences.

### Neutralizing activity of plasma

A pseudovirus-based neutralization assay was conducted using HIV reporter pseudoviruses that express the SARS-CoV-2 and BA.2.86 S proteins, along with Luciferase, as described previously(54). The assay was carried out in duplicate. Briefly, 200 TCID50 pseudoviruses were preincubated with heat-inactivated plasma samples, serially diluted three-fold (1/60–1/14), at 37 °C for 1 h in Nunc 96-well cell culture plates (Thermo Fisher Scientific, USA). Subsequently, 2×10^4^ HEK293T/hACE2 cells treated with DEAE-Dextran (Sigma-Aldrich, USA) were added. After 48 h, results were measured using the EnSight Multimode Plate Reader and BriteLite Plus Luciferase reagent (PerkinElmer, USA). The values were normalized, and the ID50 (the reciprocal dilution inhibiting 50% of the infection) was calculated by plotting the log of plasma dilution versus response, and fitting it to a four-parameter equation in Prism 8.4.3 (GraphPad Software, USA).

### Detection of undiagnosed infections based on serum antibody level changes

To identify potential undiagnosed COVID-19 infections, we analyzed fold-change (FC) of antibody levels across consecutive study timepoints. The FC for each measured antibody was determined relative to the antibody level at the initial timepoint within each timepoint interval, using the formula:

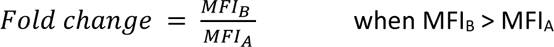

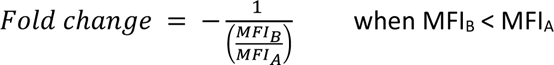

The initial FC threshold was set at 4, following the World Health Organization (WHO) recommendations(55). Two distinct approaches were applied based on whether individuals were vaccinated between the analyzed timepoints. For those vaccinated between timepoint intervals, an individual was considered infected if the FC was > 4 for IgG or IgA against the N antigen. For those not vaccinated between timepoint intervals, IgG and IgA against N and S antigens were used and an iterative tuning process was employed based on the number of diagnosed/confirmed infections detected at the threshold for each interval. Given the absence of data on confirmed non-infected individuals, threshold lowering was kept to a minimum to detect at least 75% of the diagnosed infections while minimizing the risk of false positives (**Suppl Table 1**). For the T1–T2 interval up to the T7–T8 interval, an individual was considered infected if at least two different IgG or IgA antibodies against N or S antigens exhibited a FC > 4. In later intervals, an individual was considered infected if at least one of the measured IgG or IgA antibodies had a FC > 3. A comparison of antibody level FC between putative and diagnosed infections is shown in **Suppl Fig 5**. Individuals with a positive test < 5 days before timepoint A were not considered for infection detection in the T_A_-T_B_ interval. Those vaccinated < 6 days before timepoint A were also not considered infected unless they exhibited a FC > 4 for either IgG or IgA against N. Among both vaccinated and non-vaccinated individuals between timepoint intervals, those seronegative at timepoint B were excluded from infection detection as well. In this study, we referred to diagnosed infections as “symptomatic” and undiagnosed infections, as “asymptomatic”, based on the assumption that HCW would likely seek testing if symptoms were present.

### Statistical analysis

MFI values were log_10_-transformed for analysis. Univariable and multivariable linear regression models were fitted to assess factors associated with antibody responses to SARS-CoV-2 at T9, T10 and T11. The regression coefficients (*β*) obtained from each model were converted into percentage values to facilitate interpretation. The transformed *β* value (%) was calculated using the formula ((10^*β*)-1) * 100. This indicates the percentage difference in the dependent variable associated with a 1-unit increase in the corresponding independent variable (for continuous variables) or the percentage difference in the dependent variable between the reference group and the study group (for categorical variables). Variables for adjustment for each model are specified on the Figure or Table legends.

To investigate the effect of antibody levels on the risk of SARS-CoV-2 breakthrough infection among individuals with a confirmed infection date (referred to as symptomatic along the study), we conducted a survival analysis using Kaplan–Meier estimates and multivariable Cox regression modeling. Vaccinated individuals were considered at risk from the initial timepoint (T8, T9, or T10) until the first reported episode of SARS-CoV-2 breakthrough infection, receipt of an additional vaccine dose, or the last day of study follow-up (T9, T10, or T11). Participants with asymptomatic infections (determined by serology) during the follow-up period were excluded from the analysis, as the timing of those infections could not be ascertained. The Cox proportional hazards assumption was evaluated by examining Schoenfeld residuals. To assess whether the protective effect of antibody levels varied across the three different periods (T8– T9, T9–T10, and T10–T11), we extended the Cox regression model to allow for different baseline hazard functions for each period.

To elucidate the effect of antibody levels on the risk of SARS-CoV-2 breakthrough infection in individuals with either symptomatic (with a known infection date) or asymptomatic (no known infection date) breakthrough infection events, we performed multivariable logistic regression modeling. In these analyses, vaccinated individuals were considered infected during the follow-up period (T8–T9, T9–T10, or T10–T11) if they experienced an infection without subsequent vaccination during the same period or were infected prior to receiving an additional vaccine dose (for those with diagnosed infections). Conversely, participants who did not experience an infection during the follow-up period were considered uninfected. Participants were excluded from the analysis if they were uninfected and vaccinated during the follow-up period, had an asymptomatic infection and were vaccinated during the same follow-up period, or were diagnosed infected after receiving a vaccine dose within the same period.

Missing data were handled by excluding cases with incomplete information. The sample size for each analysis is indicated in the corresponding Table and Figure legend. P-values < 0.05 were considered statistically significant. We performed the statistical analysis in R version 4.2.2.

### Study approval

The study protocols were approved by the IRB Comitè Ètic d’Investigació Clínica IDIAP Jordi Gol (code 20/162-PCV), and written informed consent was obtained from all study participants before enrollment.

### Data availability

All data produced in the present study are available upon reasonable request to the authors. The raw identifying data are protected and are not available due to data privacy laws.

## Author contributions

G.M. A.R-C, J.V-A, and C.D. designed the cohort study. A.R-M, J.V-A and A.R-C recruited participants, collected data and obtained samples. A.J., M.V., D.B., M.C., R.R., I.C. processed the samples, developed and/or performed the antibody binding assays and data preprocessing. E.P., B.T. and J. B performed the antibody neutralization analyses. R.A. contributed to design and the critical interpretation of the results. L.I, R.R., L.M-A and P.S produced the antigens. C.M.P. analyzed the data. G.M., R.A. and C.D. supervised the antibody assays and data analyses. C.M.P. wrote the first draft of the paper and R.A., G.M. and C.D reviewed the manuscript. All authors reviewed and approved the final version as submitted to the journal.

## Acknowledgments

We thank the participation of HCWs who are committed to this study and were key personnel facing the pandemic. We are grateful to Eduard Velasco at IDIAP JG for the administrative support and the nursing team for their assistance with sample collection. We also thank Selena Alonso, Natalia Diaz, Robert Mitchell, Chenjerai Jairoce, Miriam Ramirez and Laura Puyol at ISGlobal for laboratory support; Hugo Rozas for data management; Pau Serra and Daniel Parras (IDIBAPS), Carlo Carolis and Natalia Rodrigo Melero (CRG) for antigen procurement; Omicron RBD-encoding plasmids were donated by Hugo Mouquet (Institut Pasteur).

## References

1. Tseng HF, et al. Effectiveness of mRNA-1273 against SARS-CoV-2 Omicron and Delta variants. Nat Med. 2022;28(5):1063.

2. Tregoning JS, et al. Progress of the COVID-19 vaccine effort: viruses, vaccines and variants versus efficacy, effectiveness and escape. Nature Reviews Immunology 2021 21:10. 2021;21(10):626–636.

3. Lau JJ, et al. Real-world COVID-19 vaccine effectiveness against the Omicron BA.2 variant in a SARS-CoV-2 infection-naive population. Nat Med. 2023;29(2):348–357.

4. Arunachalam PS, et al. Durability of immune responses to mRNA booster vaccination against COVID-19. J Clin Invest. 2023;133(10).

5. Cao Y, et al. BA.2.12.1, BA.4 and BA.5 escape antibodies elicited by Omicron infection. Nature 2022 608:7923. 2022;608(7923):593–602.

6. Tseng HF, et al. Effectiveness of mRNA-1273 vaccination against SARS-CoV-2 omicron subvariants BA.1, BA.2, BA.2.12.1, BA.4, and BA.5. Nat Commun. 2023;14(1).

7. Planas D, et al. Distinct evolution of SARS-CoV-2 Omicron XBB and BA.2.86/JN.1 lineages combining increased fitness and antibody evasion. Nature Communications 2024 15:1. 2024;15(1):1–17.

8. Uraki R, et al. Humoral immune evasion of the omicron subvariants BQ.1.1 and XBB. Lancet Infect Dis. 2023;23(1):30–32.

9. Qu P, et al. Enhanced evasion of neutralizing antibody response by Omicron XBB.1.5, CH.1.1, and CA.3.1 variants. Cell Rep. 2023;42(5).

10. Goldblatt D, et al. Towards a population-based threshold of protection for COVID-19 vaccines. Vaccine. 2022;40(2):306–315.

11. Feng S, et al. Correlates of protection against symptomatic and asymptomatic SARS-CoV-2 infection. Nature Medicine 2021 27:11. 2021;27(11):2032–2040.

12. Gilbert PB, et al. Immune correlates analysis of the mRNA-1273 COVID-19 vaccine efficacy clinical trial. Science (1979). 2022;375(6576):43–50.

13. Wei J, et al. Antibody responses and correlates of protection in the general population after two doses of the ChAdOx1 or BNT162b2 vaccines. Nature Medicine 2022 28:5. 2022;28(5):1072–1082.

14. Martín Pérez C, et al. Correlates of protection and determinants of SARS-CoV-2 breakthrough infections 1 year after third dose vaccination. BMC Med. 2024;22(1).

15. Gilboa M, et al. Factors Associated With Protection From SARS-CoV-2 Omicron Variant Infection and Disease Among Vaccinated Health Care Workers in Israel. JAMA Netw Open. 2023;6(5):e2314757–e2314757.

16. Baerends EAM, et al. SARS-CoV-2 vaccine-induced antibodies protect against Omicron breakthrough infection. iScience. 2023;26(9):107621.

17. Perez-Saez J, et al. Long term anti-SARS-CoV-2 antibody kinetics and correlate of protection against Omicron BA.1/BA.2 infection. Nat Commun. 2023;14(1):3032.

18. Wei J, et al. Protection against SARS-CoV-2 Omicron BA.4/5 variant following booster vaccination or breakthrough infection in the UK. Nature Communications 2023 14:1. 2023;14(1):1–15.

19. Stærke NB, et al. Levels of SARS-CoV-2 antibodies among fully vaccinated individuals with Delta or Omicron variant breakthrough infections. Nat Commun. 2022;13(1).

20. Santoro A, et al. SARS-CoV-2 Breakthrough Infections According to the Immune Response Elicited after mRNA Third Dose Vaccination in COVID-19-Naïve Hospital Personnel. Biomedicines 2023, Vol 11, Page 1247. 2023;11(5):1247.

21. Dimeglio C, et al. Antibody Titers and Protection against Omicron (BA.1 and BA.2) SARS-CoV-2 Infection. Vaccines 2022, Vol 10, Page 1548. 2022;10(9):1548.

22. Walmsley S, et al. Declining Levels of Neutralizing Antibodies to SARS-CoV-2 Omicron Variants Are Enhanced by Hybrid Immunity and Original/Omicron Bivalent Vaccination. Vaccines (Basel). 2024;12(6):564.

23. Walmsley S, et al. Predictors of Breakthrough SARS-CoV-2 Infection after Vaccination. Vaccines 2024, Vol 12, Page 36. 2023;12(1):36.

24. Yamamoto S, et al. Correlates of Nucleocapsid Antibodies and a Combination of Spike and Nucleocapsid Antibodies Against Protection of SARS-CoV-2 Infection During the Omicron XBB.1.16/EG.5–Predominant Wave. Open Forum Infect Dis. 2024;11(9).

25. Canetti M, et al. Risk factors and correlates of protection against XBB SARS-CoV-2 infection among health care workers. Vaccine. 2024;42(26):126308.

26. Wajnberg A, et al. Robust neutralizing antibodies to SARS-CoV-2 infection persist for months. Science (1979). 2020;370(6521):1227–1230.

27. Rockstroh A, et al. Correlation of humoral immune responses to different SARS-CoV-2 antigens with virus neutralizing antibodies and symptomatic severity in a German COVID-19 cohort. Emerg Microbes Infect. 2021;10(1):774–781.

28. Guiomar R, et al. Monitoring of SARS-CoV-2 Specific Antibodies after Vaccination. Vaccines (Basel). 2022;10(2):154.

29. Fu J, et al. Correlation of Binding and Neutralizing Antibodies against SARS-CoV-2 Omicron Variant in Infection-Naïve and Convalescent BNT162b2 Recipients. Vaccines 2022, Vol 10, Page 1904. 2022;10(11):1904.

30. Chowdhury SH, et al. Correlation of SARS-CoV-2 Neutralization with Antibody Levels in Vaccinated Individuals. Viruses. 2023;15(3).

31. Takheaw N, et al. Correlation Analysis of Anti-SARS-CoV-2 RBD IgG and Neutralizing Antibody against SARS-CoV-2 Omicron Variants after Vaccination. Diagnostics 2022, Vol 12, Page 1315. 2022;12(6):1315.

32. Servellita V, et al. Neutralizing immunity in vaccine breakthrough infections from the SARS-CoV-2 Omicron and Delta variants. Cell. 2022;185(9):1539–1548.e5.

33. Curlin ME, et al. Omicron neutralizing antibody response following booster vaccination compared with breakthrough infection. Med. 2022;3(12):827–837.e3.

34. Lustig Y, et al. SARS-CoV-2 IgG Levels as Predictors of XBB Variant Neutralization, Israel, 2022 and 2023. Emerg Infect Dis. 2024;30(5):1050–1052.

35. Suntronwong N, et al. Strong Correlations between the Binding Antibodies against Wild-Type and Neutralizing Antibodies against Omicron BA.1 and BA.2 Variants of SARS-CoV-2 in Individuals Following Booster (Third-Dose) Vaccination. Diagnostics. 2022;12(8):1781.

36. Bobrovitz N, et al. Protective effectiveness of previous SARS-CoV-2 infection and hybrid immunity against the omicron variant and severe disease: a systematic review and meta-regression. Lancet Infect Dis. 2023;23(5):556–567.

37. Goldberg Y, et al. Protection and Waning of Natural and Hybrid Immunity to SARS-CoV-2. New England Journal of Medicine. 2022;386(23):2201–2212.

38. Srivastava K, et al. SARS-CoV-2-infection- and vaccine-induced antibody responses are long lasting with an initial waning phase followed by a stabilization phase. Immunity. 2024;57(3):587–599.e4.

39. Moncunill G, et al. Determinants of early antibody responses to COVID-19 mRNA vaccines in a cohort of exposed and naïve healthcare workers. EBioMedicine. 2022;75:103805.

40. Dobaño C, et al. High-resolution kinetics and cellular determinants of antibody response to SARS-CoV-2 over two years after COVID-19 vaccination. [published online ahead of print: January 17, 2024].

41. Altarawneh HN, et al. Effects of Previous Infection and Vaccination on Symptomatic Omicron Infections. New England Journal of Medicine. 2022;387(1):21–34.

42. Dobaño C, et al. Eleven-month longitudinal study of antibodies in SARS-CoV-2 exposed and naïve primary health care workers upon COVID-19 vaccination. Immunology. 2022;167(4):528–543.

43. Irrgang P, et al. Class switch toward noninflammatory, spike-specific IgG4 antibodies after repeated SARS-CoV-2 mRNA vaccination. Sci Immunol. 2023;8(79).

44. Lasrado N, et al. Waning immunity and IgG4 responses following bivalent mRNA boosting. Sci Adv. 2024;10(8):9945.

45. Richardson SI, et al. Antibody-dependent cellular cytotoxicity against SARS-CoV-2 Omicron sub-lineages is reduced in convalescent sera regardless of infecting variant. Cell Rep Med. 2023;4(1).

46. Imai M, et al. Efficacy of Antiviral Agents against Omicron Subvariants BQ.1.1 and XBB. New England Journal of Medicine. 2023;388(1):89–91.

47. Martín Pérez C, et al. Correlates of protection and determinants of SARS-CoV-2 breakthrough infections 1 year after third dose vaccination. BMC Med. 2024;22(1):1–16.

48. Marking U, et al. Correlates of protection and viral load trajectories in omicron breakthrough infections in triple vaccinated healthcare workers. Nature Communications 2023 14:1. 2023;14(1):1–10.

49. GISAID, via CoVariants.org [Internet]. https://ourworldindata.org/grapher/covid-variants-area?time=2021-10-25..2023-09-11&facet=none&country=~ESP. Accessed July 19, 2024.

50. Dobaño C, et al. Highly sensitive and specific multiplex antibody assays to quantify immunoglobulins M, A, and G against SARS-CoV-2 antigens. J Clin Microbiol. 2021;59(2).

51. Dobaño C, et al. Immunogenicity and crossreactivity of antibodies to the nucleocapsid protein of SARS-CoV-2: utility and limitations in seroprevalence and immunity studies. Translational Research. 2021;232:60–74.

52. Ortega N, et al. Seven-month kinetics of SARS-CoV-2 antibodies and role of pre-existing antibodies to human coronaviruses. Nature Communications 2021 12:1. 2021;12(1):1–10.

53. Planchais C, et al. Potent human broadly SARS-CoV-2-neutralizing IgA and IgG antibodies effective against Omicron BA.1 and BA.2. J Exp Med. 2022;219(7).

54. Pradenas E, et al. Stable neutralizing antibody levels 6 months after mild and severe COVID-19 episodes. Med (N Y). 2021;2(3):313–320.e4.

55. Dobaño C, et al. Antibody conversion rates to SARS-CoV-2 in saliva from children attending summer schools in Barcelona, Spain. BMC Med. 2021;19(1).

