## Supplementary Material for "Determinants of Antibody Levels and Protection against Omicron BQ.1/XBB Breakthrough Infection"

### Supplemental Material

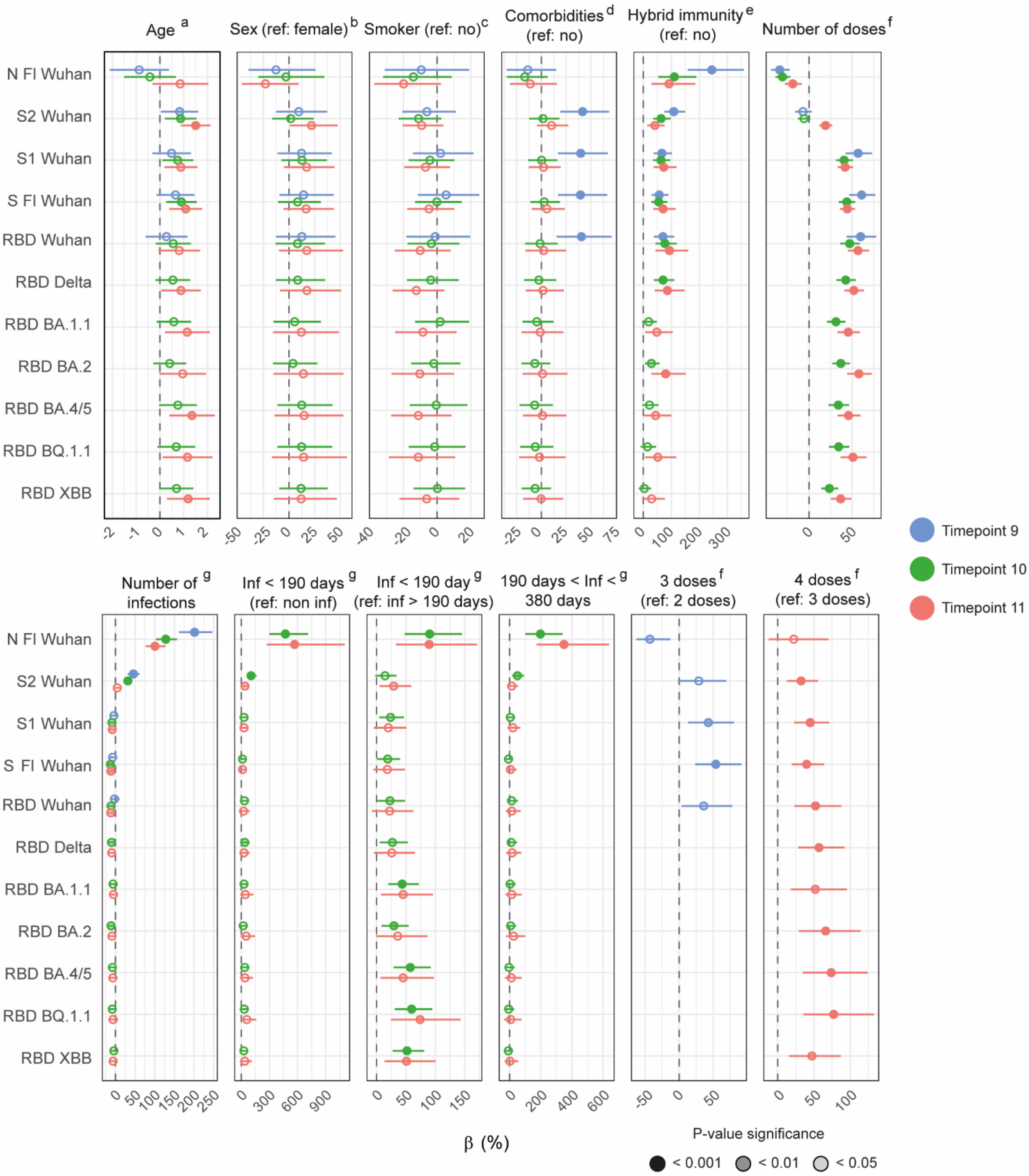


**Supplementary Figure 1. Association of clinic-demographic factors with IgG antibody levels at T9 (red), T10 (green) and T11 (blue) using univariable linear regression models in vaccinated individuals.** Beta (β) and CI values have been transformed to a percentage for an easier interpretation. The inside color of the dots represents the P value after adjustment for multiple testing by Benjamini-Hochberg, where dark color represents < 0.001, intermediate color < 0.01, light color < 0.05, and white non-significant. Full-length (Fl), Nucleocapsid (N), Receptor-binding domain (RBD), Spike (S).

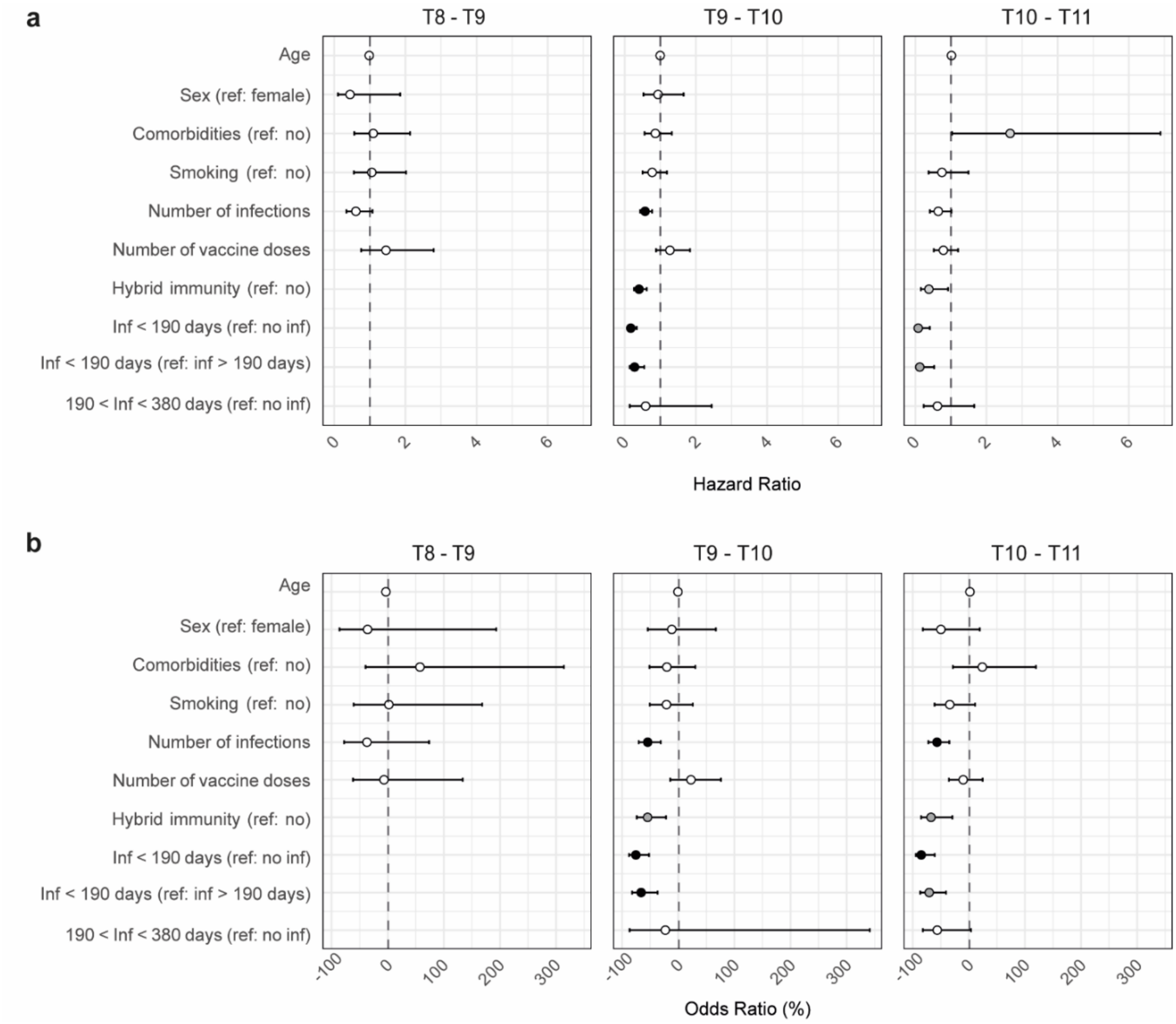


**Supplementary Figure 2.** Association of clinic-demographic factors with protection against symptomatic (**a**), and symptomatic and asymptomatic infections (**b**) using univariable Cox and Logistic regression models, respectively. In b, odds ratio and CI values have been transformed to a percentage for an easier interpretation. The color of the dots represents the P value, where black represents < 0.001, dark grey < 0.01, light grey < 0.05, and white non-significant.

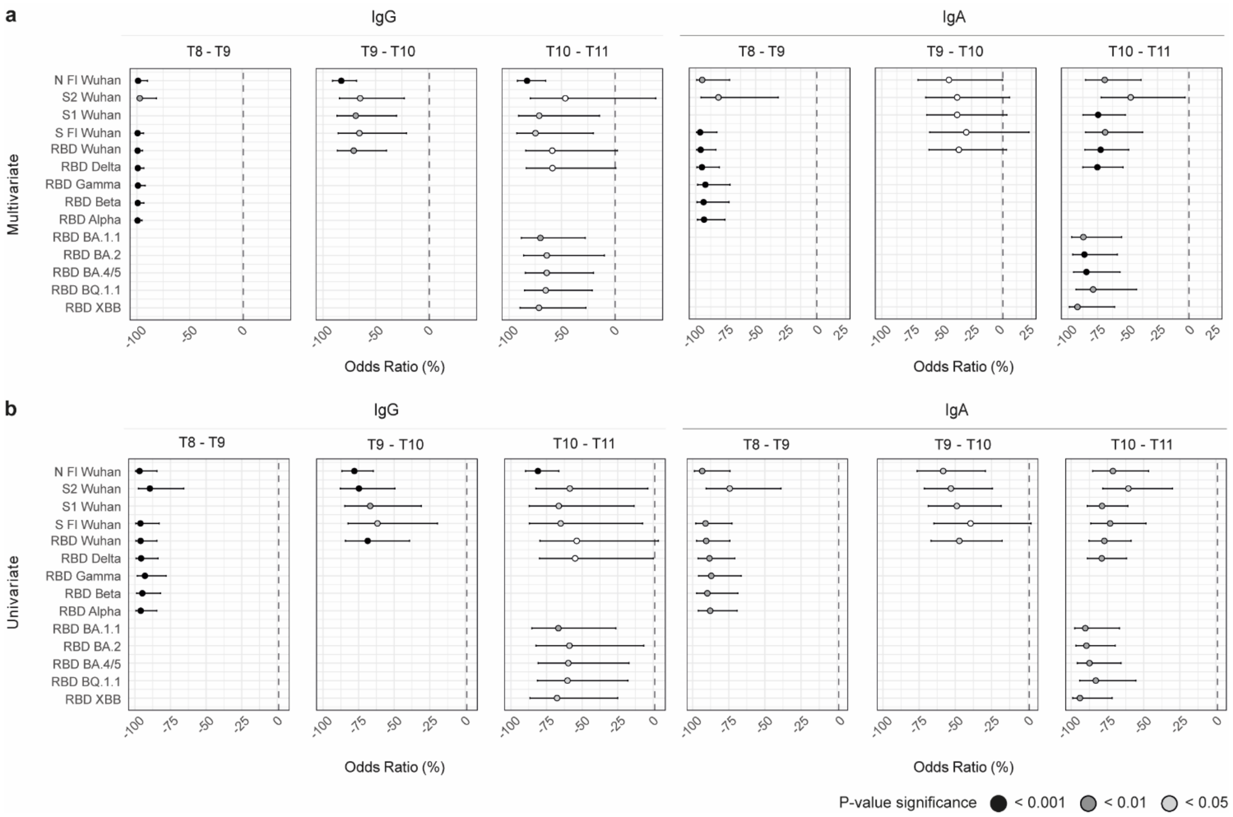


**Supplementary Figure 3. Association of antibody levels with protection against symptomatic and asymptomatic breakthrough infections in vaccinated individuals at T8 – T9, T9 – T10 and T10 – T11 periods. a**. Forest plot of multivariable Logistic Regression models. Models were adjusted by comorbidities, hybrid immunity, number of infections, number of doses, sex, smoking, and time since last exposure. **b**. Forest plot of univariable Logistic Regression models. Odds ratio and CI values have been transformed to a percentage for an easier interpretation. The color of the dots represents the P value, where black represents < 0.001, dark grey < 0.01, light grey < 0.05, and white non-significant. Full-length (Fl), Nucleocapsid (N), Receptor-binding domain (RBD), Spike (S).

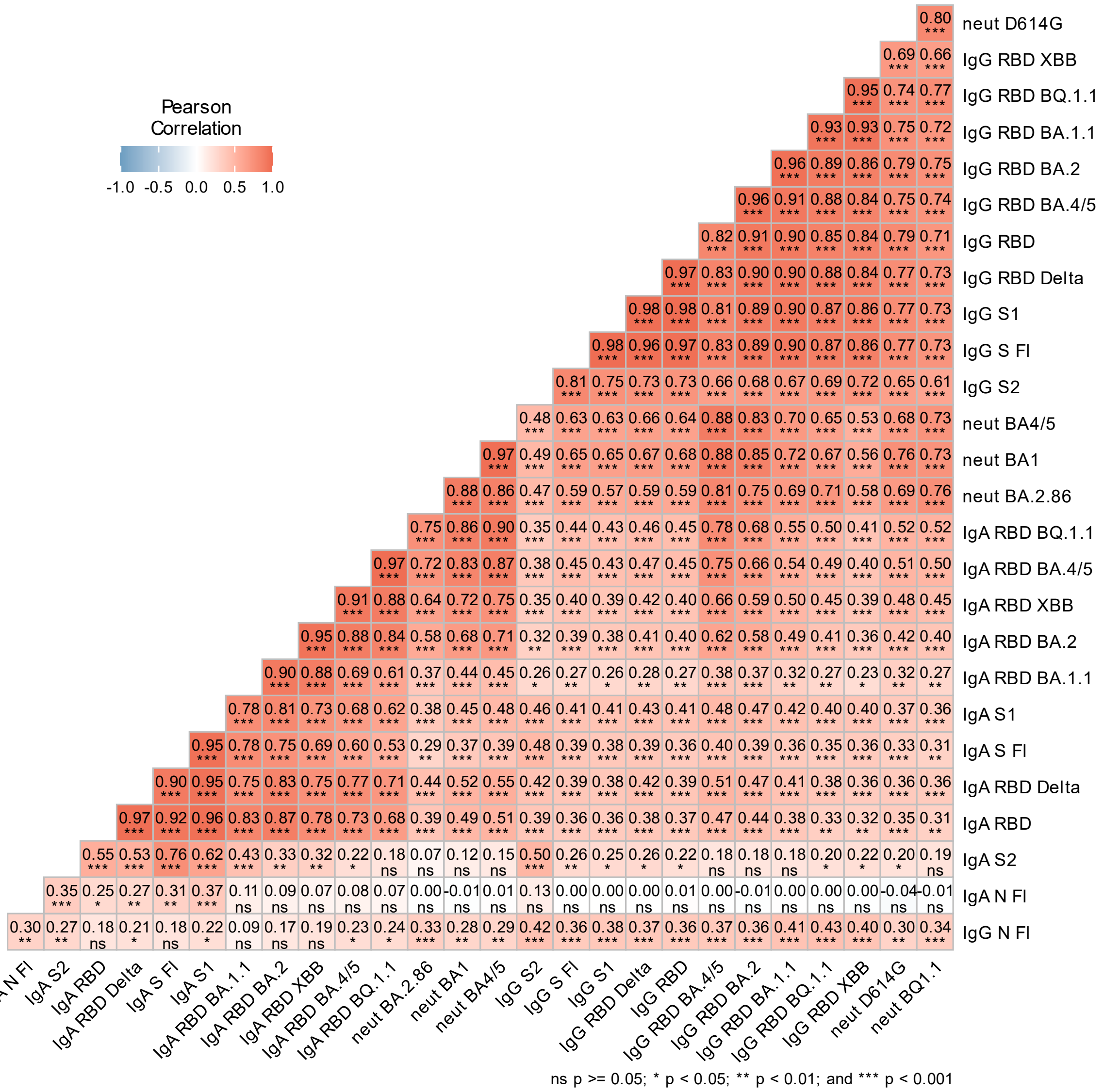


**Supplementary Figure 4.** Correlation between antibody levels and plasma neutralization capacity at T11. Pearson correlation R values are shown in the heatmap. Full-length (Fl), Nucleocapsid (N), Receptor-binding domain (RBD), Spike (S). Pearson correlation R values and correlation p-values (* p < 0.05, ** p < 0.01 , *** p < 0.001, ns) are shown in the heatmap. The D614G variant corresponds to the original Wuhan strain of the virus.

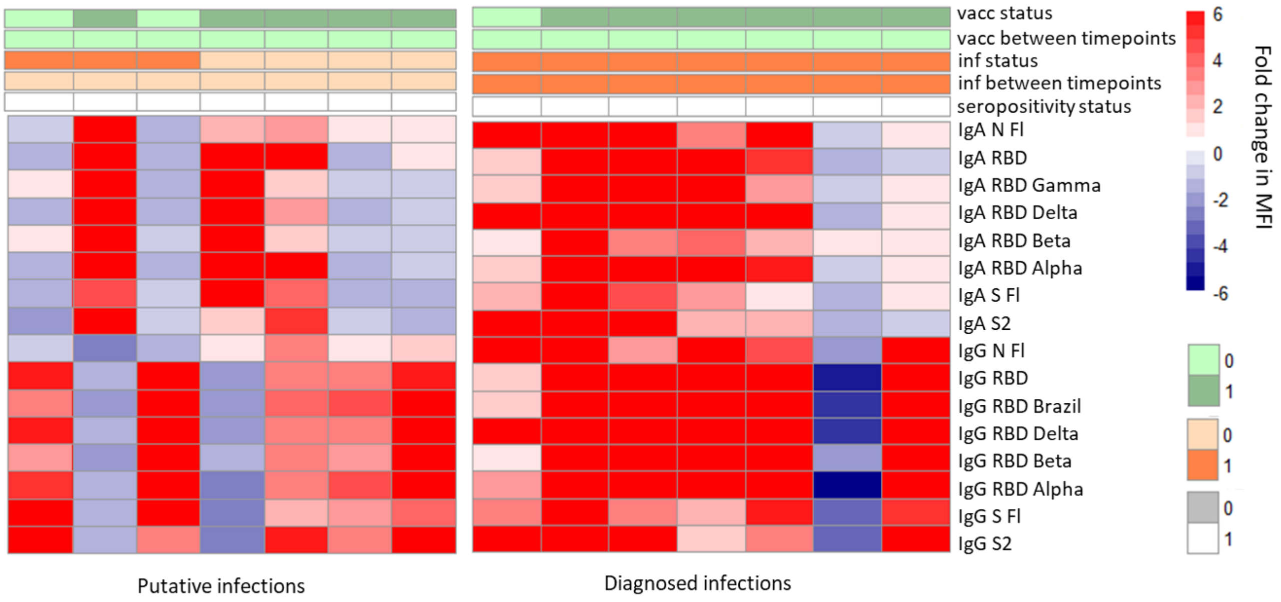


**Supplementary Figure 5. The heatmap illustrates the fold change in MFI levels of several IgG and IgA antibodies between timepoint 6 (T6) and timepoint 7 (T7) among individuals with asymptomatic putative infections and symptomatic diagnosed infections.** Each row represents a specific antibody-antigen pair, while columns represent individual samples. The color scale indicates the magnitude of fold change, increase in read and decrease in blue. Participant characteristics are overlaid above the heatmaps and include vaccination status at T7 (0 – not vaccinated, 1 – vaccinated), vaccination between T6 and T7 (0 – not vaccinated, 1 – vaccinated), infection status at T7 (0 – not infected, 1 – diagnosed as infected), infection between T6 and T7 (0 – not infected, 1 – diagnosed as infected), and seropositivity status (0 – seronegative, 1 - seropositive). Full-length (Fl), Nucleocapsid (N), Receptor-binding domain (RBD), Spike (S).

**Supplementary Table 1.** Total number of diagnosed infections, and diagnosed infections detected with antibody levels fold change analysis per timepoint interval among those not vaccinated between timepoints.

| Interval | Diagnosed infections (n) | Detected diagnosed infections (n) | Detected diagnosed infections (%) |
|---|---|---|---|
| T1 - T2 | 1 | 1 | 100 |
| T3 - T4 | 0 | 0 | - |
| T4 - T5 | 0 | 0 | - |
| T5 - T6 | 0 | 0 | - |
| T6 - T7 | 7 | 6 | 85 |
| T7 - T8 | 5 | 5 | 100 |
| T8 - T9 | 42 | 39 | 93 |
| T9 - T10 | 89 | 68 | 76 |
| T10 - T11 | 33 | 29 | 88 |

**Supplementary Table 2.** Characteristics of study participants at T9.

| | | **N or mean ± SD** |
|---|---|---|
| Age | | 48.6 ± 10,7 |
| Sex | Male | 61 |
| | Female | 316 |
| Comorbidities | Yes | 252 |
| | No | 125 |
| Smoking | Yes | 160 |
| | No | 217 |
| Vaccine doses | 0 | 15 |
| | 1 | 29 |
| | 2 | 88 |
| | 3 | 245 |
| Number of infections* | 0 | 77 |
| | 1 | 195 |
| | 2 | 95 |
| | 3 | 9 |
| | 4 | 1 |
| First dose type | Astrazeneca | 5 |
| | Janssen | 1 |
| | Moderna | 4 |
| | Pfizer | 342 |
| | NA | 10 |
| Second dose type | Astrazeneca | 4 |
| | Moderna | 14 |
| | Pfizer | 302 |
| | NA | 13 |
| Third dose type | Moderna | 118 |
| | Pfizer | 99 |
| | NA | 28 |
| First exposure type | Infection | 206 |
| | Vaccination | 171 |

* Includes symptomatic and asymptomatic infections.

**Supplementary Table 3.** Characteristics of study participants at T10.

| | | N or mean ± SD |
|---|---|---|
| Age | | 50 ± 10,7 |
| Sex | Male | 58 |
| | Female | 334 |
| Comorbidities | Yes | 260 |
| | No | 132 |
| Smoking | Yes | 166 |
| | No | 226 |
| Vaccine doses | 0 | 16 |
| | 1 | 26 |
| | 2 | 86 |
| | 3 | 223 |
| | 4 | 41 |
| Number of infections* | 0 | 43 |
| | 1 | 174 |
| | 2 | 141 |
| | 3 | 32 |
| | 4 | 2 |
| First dose type | Astrazeneca | 5 |
| | Janssen | 1 |
| | Moderna | 4 |
| | Pfizer | 324 |
| | NA | 10 |
| Second dose type | Astrazeneca | 4 |
| | Moderna | 14 |
| | Pfizer | 291 |
| | NA | 12 |
| Third dose type | Moderna | 109 |
| | Pfizer | 102 |
| | NA | 28 |
| Fourth dose type | Pfizer | 37 |
| | NA | 4 |
| First exposure type | Infection | 208 |
| | Vaccination | 184 |

* Includes symptomatic and asymptomatic infections

**Supplementary Table 4.** Summary of p-values assessing the interaction between time period and IgG or IgA antibody levels on protection against symptomatic and asymptomatic infections using multivariable Logistic regression models. Models were adjusted by comorbidities, hybrid immunity, number of exposures, sex, smoking, and time since last exposure. Interaction terms were added between time period and hybrid immunity, time period and number of exposures, time period and time since the last exposure and time period and antibody levels. P values were adjusted for multiple testing by Benjamini-Hochberg method. Full-length (Fl), Nucleocapsid (N), Receptor-binding domain (RBD), Spike (S).

| Timepoint interval | Isotype | Analyte | P-value Interaction | Adjusted Interaction P-value |
|---|---|---|---|---|
| T8 – T9 vs T9 – T10 | IgG | N Fl | 0.045 | 0.079 |
| T8 – T9 vs T9 – T10 | IgG | RBD | 0.003 | 0.013 |
| T8 – T9 vs T9 – T10 | IgG | S Fl | 0.004 | 0.013 |
| T8 – T9 vs T9 – T10 | IgG | S2 | 0.051 | 0.080 |
| T8 – T9 vs T10 – T11 | IgG | N Fl | 0.024 | 0.048 |
| T8 – T9 vs T10 – T11 | IgG | RBD | < 0.001 | 0.006 |
| T8 – T9 vs T10 – T11 | IgG | RBD Delta | < 0.001 | 0.006 |
| T8 – T9 vs T10 – T11 | IgG | S Fl | 0.005 | 0.015 |
| T8 – T9 vs T10 – T11 | IgG | S2 | 0.017 | 0.042 |
| T9 – T10 vs T10 – T11 | IgG | N Fl | 0.920 | 0.991 |
| T9 – T10 vs T10 – T11 | IgG | RBD | 0.487 | 0.621 |
| T9 – T10 vs T10 – T11 | IgG | S Fl | 0.806 | 0.941 |
| T9 – T10 vs T10 – T11 | IgG | S1 | 0.998 | 0.998 |
| T9 – T10 vs T10 – T11 | IgG | S2 | 0.382 | 0.535 |
| T8 – T9 vs T9 – T10 | IgA | N Fl | 0.017 | 0.079 |
| T8 – T9 vs T9 – T10 | IgA | RBD | 0.001 | 0.013 |
| T8 – T9 vs T9 – T10 | IgA | S | 0.002 | 0.013 |
| T8 – T9 vs T9 – T10 | IgA | S2 | 0.133 | 0.169 |
| T8 – T9 vs T10 – T11 | IgA | N Fl | 0.066 | 0.104 |
| T8 – T9 vs T10 – T11 | IgA | RBD | 0.048 | 0.097 |
| T8 – T9 vs T10 – T11 | IgA | RBD Delta | 0.109 | 0.153 |
| T8 – T9 vs T10 – T11 | IgA | S Fl | 0.041 | 0.096 |
| T8 – T9 vs T10 – T11 | IgA | S2 | 0.240 | 0.259 |
| T9 – T10 vs T10 – T11 | IgA | N Fl | 0.240 | 0.259 |
| T9 – T10 vs T10 – T11 | IgA | RBD | 0.028 | 0.079 |
| T9 – T10 vs T10 – T11 | IgA | S Fl | 0.067 | 0.1044 |
| T9 – T10 vs T10 – T11 | IgA | S1 | 0.027 | 0.079 |
| T9 – T10 vs T10 – T11 | IgA | S2 | 0.674 | 0.674 |

**Supplementary Table 5.** Summary of p-values assessing the interaction between time period and IgG or IgA antibody levels on protection against symptomatic infections using multivariable Cox regression models. Models were adjusted by comorbidities, hybrid immunity, number of exposures, sex, smoking, and time since last exposure. Interaction terms were added between time period and hybrid immunity, time period and number of exposures, time period and time since the last exposure and time period and antibody levels. P values were adjusted for multiple testing by Benjamini-Hochberg method. Full-length (Fl), Nucleocapsid (N), Receptor-binding domain (RBD), Spike (S).

| Timepoint interval | Isotype | Analyte | P-value Interaction | Adjusted Interaction P-value |
|---|---|---|---|---|
| T8 – T9 vs T9 – T10 | IgG | N Fl | 0.008 | 0.033 |
| T8 – T9 vs T9 – T10 | IgG | RBD | < 0.001 | 0.004 |
| T8 – T9 vs T9 – T10 | IgG | S Fl | < 0.001 | 0.004 |
| T8 – T9 vs T9 – T10 | IgG | S2 | 0.009 | 0.033 |
| T8 – T9 vs T10 – T11 | IgG | N Fl | 0.012 | 0.035 |
| T8 – T9 vs T10 – T11 | IgG | RBD | 0.015 | 0.035 |
| T8 – T9 vs T10 – T11 | IgG | RBD Delta | 0.025 | 0.046 |
| T8 – T9 vs T10 – T11 | IgG | S Fl | 0.075 | 0.118 |
| T8 – T9 vs T10 – T11 | IgG | S2 | 0.026 | 0.046 |
| T9 – T10 vs T10 – T11 | IgG | N Fl | 0.804 | 0.866 |
| T9 – T10 vs T10 – T11 | IgG | RBD | 0.723 | 0.843 |
| T9 – T10 vs T10 – T11 | IgG | S Fl | 0.294 | 0.412 |
| T9 – T10 vs T10 – T11 | IgG | S1 | 0.368 | 0.468 |
| T9 – T10 vs T10 – T11 | IgG | S2 | 0.872 | 0.872 |
| T8 – T9 vs T9 – T10 | IgA | N Fl | 0.148 | 0.245 |
| T8 – T9 vs T9 – T10 | IgA | RBD | < 0.001 | 0.004 |
| T8 – T9 vs T9 – T10 | IgA | S Fl | < 0.001 | 0.006 |
| T8 – T9 vs T9 – T10 | IgA | S2 | 0.010 | 0.050 |
| T8 – T9 vs T10 – T11 | IgA | N Fl | 0.473 | 0.473 |
| T8 – T9 vs T10 – T11 | IgA | RBD | 0.044 | 0.156 |
| T8 – T9 vs T10 – T11 | IgA | RBD Delta | 0.175 | 0.245 |
| T8 – T9 vs T10 – T11 | IgA | S Fl | 0.110 | 0.221 |
| T8 – T9 vs T10 – T11 | IgA | S2 | 0.265 | 0.309 |
| T9 – T10 vs T10 – T11 | IgA | N Fl | 0.383 | 0.412 |
| T9 – T10 vs T10 – T11 | IgA | RBD | 0.158 | 0.245 |
| T9 – T10 vs T10 – T11 | IgA | S Fl | 0.107 | 0.221 |
| T9 – T10 vs T10 – T11 | IgA | S1 | 0.094 | 0.221 |
| T9 – T10 vs T10 – T11 | IgA | S2 | 0.192 | 0.245 |

**Supplementary Table 6.** Summary of p-values assessing the interaction between time period and IgG or IgA antibody levels on protection against symptomatic, and symptomatic and asymptomatic infections using multivariable Cox (**a**) or Logistic regression models (**b**), respectively. Anti-Wuhan RBD antibodies were selected for T8 – T9 period, while anti-Omicron RBD antibodies were selected for T10 – T11 period. Models were adjusted by comorbidities, hybrid immunity, number of exposures, sex, smoking, and time since last exposure. Interaction terms were added between time period and hybrid immunity, time period and number of exposures, time period and time since the last exposure, and time period and antibody levels. Full-length (Fl), Nucleocapsid (N), Receptor-binding domain (RBD), Spike (S).

a

| Timepoint interval | Isotype | Analyte at T8 | Analyte at T10 | Interaction P-value |
|---|---|---|---|---|
| T8 – T9 vs T10 – T11 | IgG | RBD Wuhan | RBD BA.1.1 | 0.084 |
| | IgG | | RBD BA.2 | 0.081 |
| | IgG | | RBD BA.4/5 | 0.053 |
| | IgG | | RBD BQ.1.1 | 0.040 |
| | IgG | | RBD XBB | 0.230 |
| | IgA | | RBD BA.1.1 | 0.408 |
| | IgA | | RBD BA.2 | 0.643 |
| | IgA | | RBD BA.4/5 | 0.820 |
| | IgA | | RBD BQ.1.1 | 0.595 |
| | IgA | | RBD XBB | 0.481 |

b

| Timepoint interval | Isotype | Analyte at T8 | Analyte at T10 | Interaction P-value |
|---|---|---|---|---|
| T8 – T9 vs T10 – T11 | IgG | RBD Wuhan | RBD BA.1.1 | 0.003 |
| | IgG | | RBD BA.2 | 0.001 |
| | IgG | | RBD BA.4/5 | 0.001 |
| | IgG | | RBD BQ.1.1 | 0.002 |
| | IgG | | RBD XBB | 0.003 |
| | IgA | | RBD BA.1.1 | 0.384 |
| | IgA | | RBD BA.2 | 0.338 |
| | IgA | | RBD BA.4/5 | 0.241 |
| | IgA | | RBD BQ.1.1 | 0.122 |
| | IgA | | RBD XBB | 0.690 |

**Supplementary Table 7. a**. Summary of multivariable Cox regression models assessing the association between plasma neutralizing capacity at T9 and protection against symptomatic infections over the T9 – T10 period. **b**. Summary of multivariable Logistic regression models assessing the association between plasma neutralizing capacity at T9 and protection against symptomatic and asymptomatic infections over the T9 – T10 period. The D614G variant corresponds to the original Wuhan strain of the virus. Odds ratio and CI values have been transformed to a percentage for an easier interpretation.

a

| Analyte | HR | Upper CI | Lower CI | P-value |
|---|---|---|---|---|
| D614G | 0.25 | 0.53 | 0.12 | < 0.001 |
| BA.1 | 0.24 | 0.52 | 0.11 | < 0.001 |
| BA.4.5 | 0.20 | 0.50 | 0.08 | < 0.001 |
| BQ.1.1 | 0.12 | 0.51 | 0.03 | 0.004 |

b

| Analyte | OR (%) | Upper CI | Lower CI | P-value |
|---|---|---|---|---|
| D614G | -83.2 | -57.3 | -94.2 | < 0.001 |
| BA.1 | -82.6 | -57.7 | -93.6 | < 0.001 |
| BA.4.5 | -85.8 | -60.3 | -95.7 | < 0.001 |
| BQ.1.1 | -91.7 | -63.6 | -98.6 | 0.003 |
